# The effect of a specialist paramedic primary care rotation on appropriate non-conveyance decisions (SPRAINED) study: a controlled interrupted time series analysis

**DOI:** 10.1101/2020.08.06.20169334

**Authors:** Richard Pilbery, Tracey Young, Andrew Hodge

## Abstract

**Introduction:** NHS ambulance service conveyance rates in the UK are almost 70%, despite an increase in non-emergency cases. This is increasing the demands on crowded emergency departments (ED) and contributes to increased ambulance turnaround times. Yorkshire Ambulance Service introduced a specialist paramedic (SP) role to try and address this, but non-conveyance rates in this group have not been as high as expected.

**Methods:** We conducted a controlled interrupted time series analysis using data from incidents between June 2017 and December 2019, to study appropriate non-conveyance rates before and after a GP placement. A costing analysis examined the average cost per appropriate non-conveyance achieved for patients receiving care from intervention group SPs pre- and post-placement was also conducted.

**Results:** 7349 incidents attended by intervention group SPs were eligible for inclusion. Following removal of cases with missing data, 5537/7349 (75.3%) cases remained. Post-placement, the intervention group demonstrated an increase in appropriate non-conveyance rate of 35.0% (95%CI 23.8–46.2%, p<0.001), and a reduction in the trend of appropriate non-conveyance of −1.2% (95%CI −2.8–0.5%, p=0.156), relative to the control group.

Post-placement, the cost per appropriate non-conveyance for intervention group SPs was a mean of £509.38 (95% bootstrapped CI £455.32–£564.59) versus £1258.04 (95% bootstrapped CI £1232.64–£1284.04) for the same group in the pre-placement phase. This represents a mean saving of £748.66 per appropriate non-conveyance (95% bootstrapped CI £719.45–£777.32) and a cost-effectiveness ratio of £2141.15 per percentage increase in appropriate non-conveyance (95% bootstrapped CI £2057.62–£2223.12).

**Conclusion:** In this single UK NHS ambulance service study, we found a clinically important and statistically significant increase in appropriate non-conveyance rates by specialist paramedics who had completed a 10-week GP placement. This improvement persisted for the 12-month period following the placement and demonstrated cost savings compared to usual care.

**What this study adds:** *What is known about this subject:*

- UK ambulance service conveyance rates are almost 70% despite an increase in the number of non-emergency cases
- Health Education England funded a pilot in 2018 to rotate paramedics into a range of health settings to improve patient care and relieve pressures on primary care services.

*What this study adds:*

- Clinically important and statistically significant increases in appropriate non-conveyance rates can be achieved by specialist paramedics who complete a 10-week primary care placement.
- This improvement is sustained for at least 12-months following the placement cost-effective.

## Introduction

The National Health Service (NHS) in the United Kingdom is facing a 5% year-on-year increase in demand for urgent and emergency care services [1]. In 2018/19, ambulance services in England provided a face-to-face assessment to nearly 7.9 million incidents, of which 5.4 million were conveyed to hospital [2]. This non-conveyance rate of nearly 70% is occurring despite an increase in non-emergency cases and continues to place increasing demands on already crowded emergency departments (EDs), leading to decreased availability of ambulances as turnaround times at hospitals increase [3]. ED overcrowding is a significant issue for patients, resulting in poorer quality of care, increased healthcare costs and potentially, increased mortality [4–7].

Yorkshire Ambulance Service NHS Trust (YAS) have been early adopters of initiatives to respond appropriately to the increase in non-emergency cases, with the introduction of advanced paramedics working in the role of Emergency Care Practitioners (ECP) since 2004 [8]. Even without selective dispatching, ECPs consistently have non-conveyance rates double that of other paramedics in the Trust. Since 2015, the specialist paramedic (SP) role has been introduced in YAS, with education comprising of a 1 year university program. However, during the time period of this study (2017–2019), Trust-wide YAS SP non-conveyance rates were around 37.5%, compared to the overall Trust non-conveyance rate of 30%. In contrast, ECPs had non-conveyance rates of over 59% for all call categories.

In 2018, Health Education England (HEE) funded a pilot scheme to rotate paramedics into a range of healthcare settings, with the aim of improving patient care and relieving pressures on primary care, ambulance services and other parts of the NHS in a sustainable way [9]. A subsequent economic evaluation estimated that the rotating paramedics could save in the region of £275,000 per year in avoidable conveyance and subsequent admission to hospital compared to historic controls. However, data from Yorkshire included 5 ECPs who participated in the pilot and the analysis did not adjust for the difference in patient acuity between the pre- and post-placement phases [10].

The HEE pilot also presented an opportunity to further develop the decision-making of SPs, with the potential to deliver patient and cost-benefits that were anticipated when the role was created. This study aimed to evaluate whether a primary care placement appropriately increased the level and trend of non-conveyance decisions made by SPs following a 10-week GP placement, in a cost-effective manner.

## Objectives

The primary objective was to determine the change and trend in proportion of appropriate non-conveyance decisions by specialist paramedics who have completed a 10-week placement in a General Practitioner (GP) practice. The secondary objective was to compare the cost-effectiveness of specialist paramedics who have completed a 10-week placement in a GP practice compared to usual care pre-placement.

### Methods

This study was a natural experiment using routinely collected observational data. We utilised a controlled interrupted time series (CITS) analysis method to detect any change in the level and trend of appropriate non-conveyance decisions by SPs following a 10-week GP practice placement. The cost-effectiveness analysis examined the cost per appropriate non-conveyance achieved for patients receiving care for the same group of SPs, comparing their post-placement performance with usual care pre-placement. In addition, we calculated the cost-effectiveness ratio to determine the difference in cost between groups per percent increase in appropriate non-conveyance.

National reporting of non-conveyance rates describe the proportion of patients who call 999 and are not transported to ED. If a resource e.g. ambulance is not sent to scene, this call is classed as a ‘hear and treat’ call. However, if an ambulance is sent to scene but the patient is not subsequently transported to hospital, this is classed as a ‘see and treat’ call. SPs participating in the YAS HEE pilot, did not have the opportunity to undertake ‘hear and treat’ calls and so non-conveyance for the purpose of this study, refers only to ‘see and treat’ calls.

### Setting

YAS provides 24-hour emergency and health care services for the county of Yorkshire in the north of England and receives more than 998,500 emergency calls each year. The organisation employs approximately 1200 operational paramedics, 118 SPs and 33 ECPs. Following an internal recruitment process, 5 ECPs and 10 SPs were interviewed and seconded to work with a number of primary care organisations in Leeds. ECPs and SPs qualify with additional degree level training in clinical assessment, decision making and minor illness and injury management, and those selected for the pilot had a minimum of 2 years’ experience in these roles.

The ECPs and SPs rotated between providing a home visiting service for 15 GP surgeries for 10 weeks, which included an induction program of primary care specific conditions, having their clinical practice observed by a GP and a named GP mentor. This was followed by rotation back into front line operations, either responding to 999 calls as a solo responder in a rapid response car or working in the emergency operations centre (EOC) staffing a dedicated dispatch desk for SPs, where they identified appropriate 999 calls according to their skills. Additional equipment carried by SPs included glue and butterfly stitches (Steri-Strips) for minor wound care, and a small range of steroids and antibiotics for common illnesses or exacerbation of chronic conditions.

### Data sources

We used routinely collected computer aided dispatch (CAD) and patient record data to identify all cases attended by the 10 SPs who had completed a GP placement in the Leeds area. For operational reasons, these placements were staggered, with the first paramedics entering the rotation in June 2018 and the final paramedics completing their placements at the end of December 2018. In order to obtain sufficient data pre- and post-pilot, all cases attended by these paramedics in the 12 month period prior to the placement commencing and 12 months after the placement had completed were obtained. To count as an ‘attendance’, the SPs name had to appear on the patient record.

To take account of case-mix and paramedic experience in the pre- and post-placement phase, a matched comparison (control) group consisting of incidents in Yorkshire occurring between the 1st June 2017 to 31st December 2019 was obtained. This cohort of patients received a face-to-face assessment by paramedics and SPs who did not take part in the GP rotation. Patients seen exclusively by staff not registered with the Health and Care Professions Council (HCPC) e.g. associate ambulance practitioners and emergency medical technicians were excluded.

Since YAS does not keep a record of paramedic registration beyond the current 2-year registration cycle, it was not possible to determine how long staff had been registered as a paramedic using data from the Service. Instead, we identified when staff were first entered into the HCPC paramedic register.

### Study variables

We hypothesised that appropriate non-conveyance was likely to increase following the 10-week placement, but needed to ensure that we took account of factors previously identified as being important when pre-hospital clinicians make non-conveyance decisions [11]. To achieve this, we aimed to match the control and intervention groups on patient age, sex, clinician determined working impression, time, month and year of call, triage category, lowest recorded National Early Warning Score (NEWS) threshold, lower super output area (LSOA) rural/urban classification, index of multiple deprivation decile and proportion of population in LSOA with a long-term physical or mental illness. Finally, we included the number of years clinicians had been registered as a paramedic and their role designation at the time of the incident.

Since the ambulance service does not routinely capture outcome data for all patients, we pragmatically defined appropriate non-conveyance as any patient episode where the patient was not transferred to hospital and no further calls were made to the ambulance service in the following 72 hours.

### Matching

Matching was performed utilising a genetic algorithm and computed using the R (v3.6.0) statistics package ‘Matching’ (v4.9-7) [12]. Genetic algorithms are a subgroup of evolutionary computing which as the name suggests, imitate biological processes of reproduction and natural selection to solve according to ‘fitness’ [13]. The ‘Matching’ package uses this algorithm to find the optimal balance between groups by examining the cumulative probability distribution functions of a variety of standardised statistics such as t-tests and Kolmogorov-Smirnov tests.

Cases where the patient record could not be located or where data was missing were excluded. With the exception of ECPs (who were removed since their non-conveyance rates are consistently higher than other clinicians), it was not possible to accurately determine what role designation paramedics in the control group had. As a result, this had to be removed as a matching variable.

### Statistical methods

#### Sample size calculation

The sample size was constrained by the fact that this was an observational study with only 12 months pre- and post-placement available. Based on a previous audit of 999 call data, we anticipated that there would be approximately 700 patient episodes per month. Given that the time series analysis covered a 24 month time period in total, it was anticipated that approximately 33,600 incidents would be included in the dataset.

#### Summary of baseline data

Descriptive statistics were used to summarise the data pre- and post-placement and between intervention group SPs and the control group, to illustrate the success of matching. Median and interquartile ranges were reported for continuous variables, and counts and proportions reported for categorical data.

#### Primary outcome analysis

We conducted a retrospective analysis of appropriate non-conveyance before and after the GP placement, using segmented regression as part of a CITS design [14]. Since the SP placements were staggered, the actual month and year was not utilised. Instead, the number of months before and after the placement were used, so that month 1 was the month that occurred 12 months prior to the GP placement for all SPs and month 24, the month that occurred 12 months after the placement. It was anticipated that this would remove or reduce any autocorrelation. However, we checked for auto-regression and moving averages by performing the Durbin-Watson test and by plotting autocorrelation function and partial autocorrelation function plots. Coefficients from the model were used to predict the absolute change and trend in appropriate non-conveyance following the GP placement, relative to the control group.

#### Secondary outcome analysis

Salary costs were calculated for the 10 week GP placement and divided by the number of incidents attended by SPs to calculate a per-incident cost. SPs were assumed to be salaried at NHS Agenda for Change mid-band 6, which was £31,121 for 2018/19. Education costs were not included since all SPs had already undertaken the education component prior to the HEE pilot commencing. The resource use related to the 999 call handling, dispatch of an ambulance, cost of conveyance and admission to the ED was calculated using reference costs published by NHS Improvement for the 2018/19 financial year [15]. The reference costs are a flat rate, irrespective of the number of resources or skill mix the Trust allocates to an incident. A 999 call made to the EOC costs £7.33, an ambulance see and treat response costs £209.38, an ambulance see, treat and convey response costs £257.34 and an ED attendance costs £135.00.

A segmented regression analysis was conducted, similar to that of the primary objective, but with incident cost as the dependent variable and the addition of safe non-conveyance as an independent variable. This allowed us to adjust for any case mix differences between the pre- and post-placement phases. Bootstrapping was used to estimate uncertainty (reported as 95% bootstrapped confidence intervals) around cost estimates. Costs relating to patients seen by intervention group SPs post-placement was compared with costs arising from patients seen by the intervention group SPs pre-placement, and the results presented as the cost per appropriate conveyance and cost-effectiveness ratio.

#### Patient and public involvement

YAS patient research ambassadors were consulted with respect to the validity of conducting the study and the wording of the Plain English summary.

## Results

As part of the HEE rotational paramedic pilot, 10 SPs undertook a 10 week placement in a primary care setting in the Leeds area. Five commenced their placement in June 2018, with a further three starting in August 2018 and the final two starting their placement in October 2018.

Between 1st June 2017 and 31st December, 2019 there were 8849 incidents attended by one of the intervention group SPs. Once data was adjusted to remove any cases during the 10 week GP placement, and outside of the 12 months prior to the start of the rotation and 12 months after the end of the rotation, 7349 cases remained (Figure 1). A further 6 had no sex recorded, 15 had no age recorded, 8 had no post code and 4 cases were excluded due to either a missing index of multiple deprivation decile (3 cases), rural urban classification (3 cases) and/or prevalence of missing long-term condition data (4 cases). Finally, no working impression was included in 1785 cases, resulting in a final dataset of 5537/7349 (75.3%) cases for inclusion in the final analysis. Due to the high number of missing working impressions, a sensitivity analysis was performed excluding the working impression as a variable (Supplementary 1).

**Figure 1:**
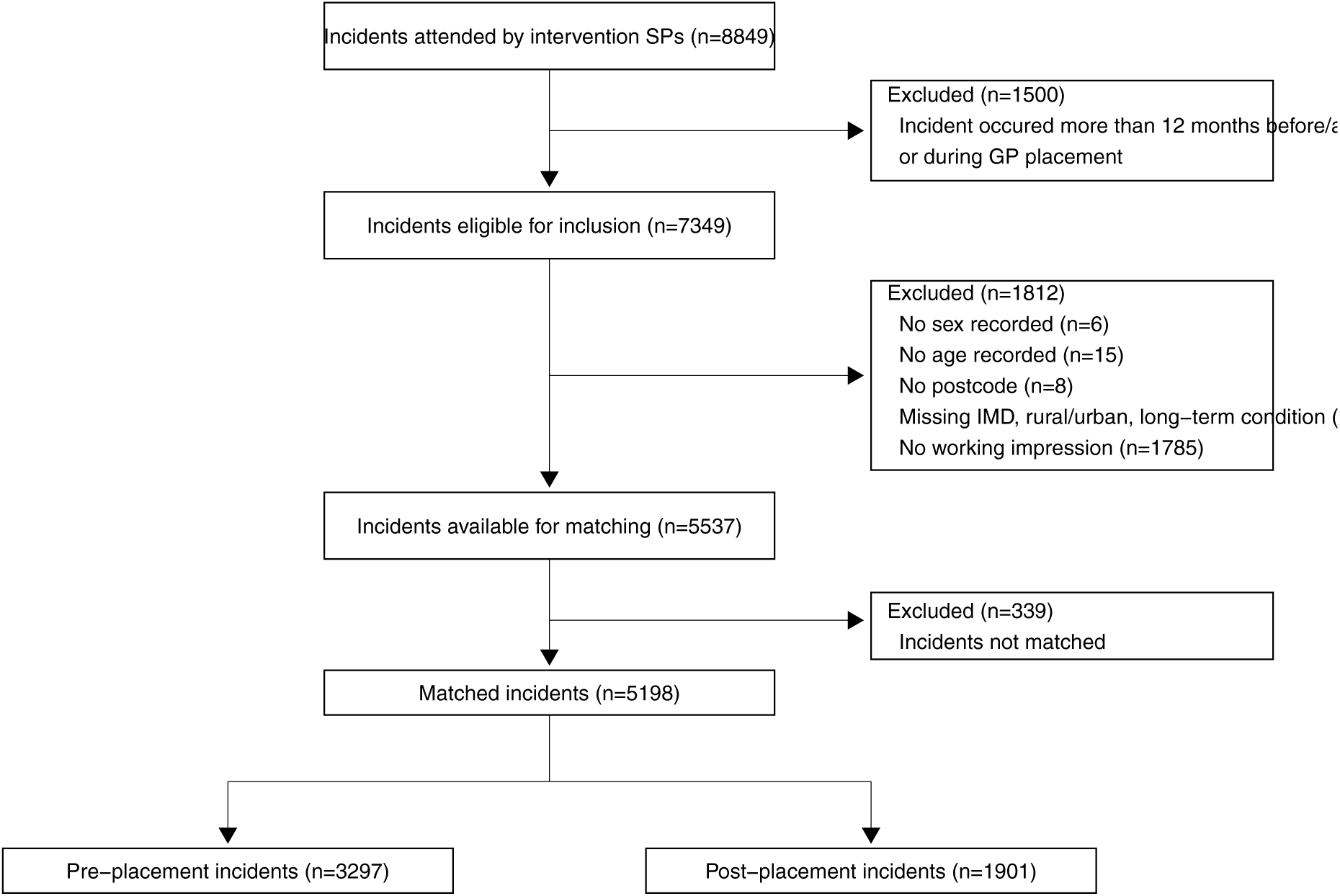
STROBE flow chart of patient selection

### Matched dataset for analysis

The matching algorithm utilised 5198/5537 (93.9%) cases (Table 1). Overall, the control group was closely matched to the rotational paramedic (intervention group) incidents (defined as less than 10% in standardised mean difference). Only the NEWS risk category and prevalence of long-term conditions were outside this limit.

**Table 1:**
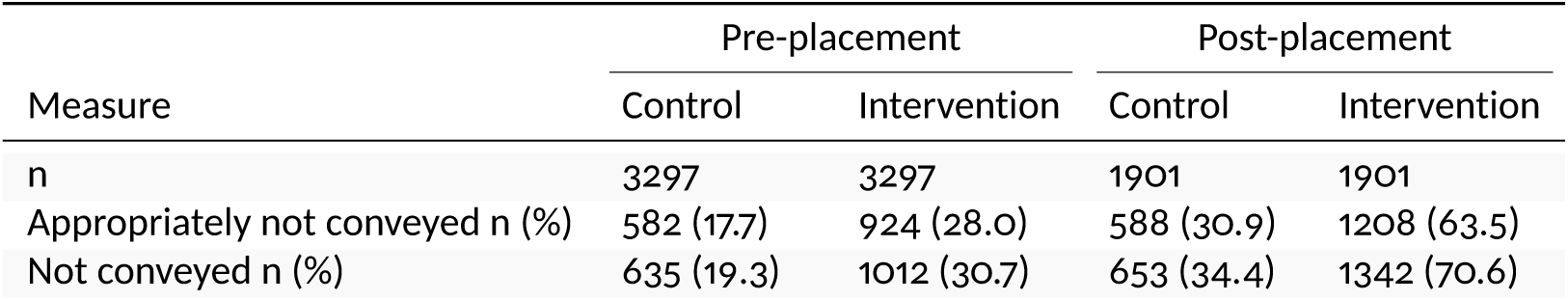
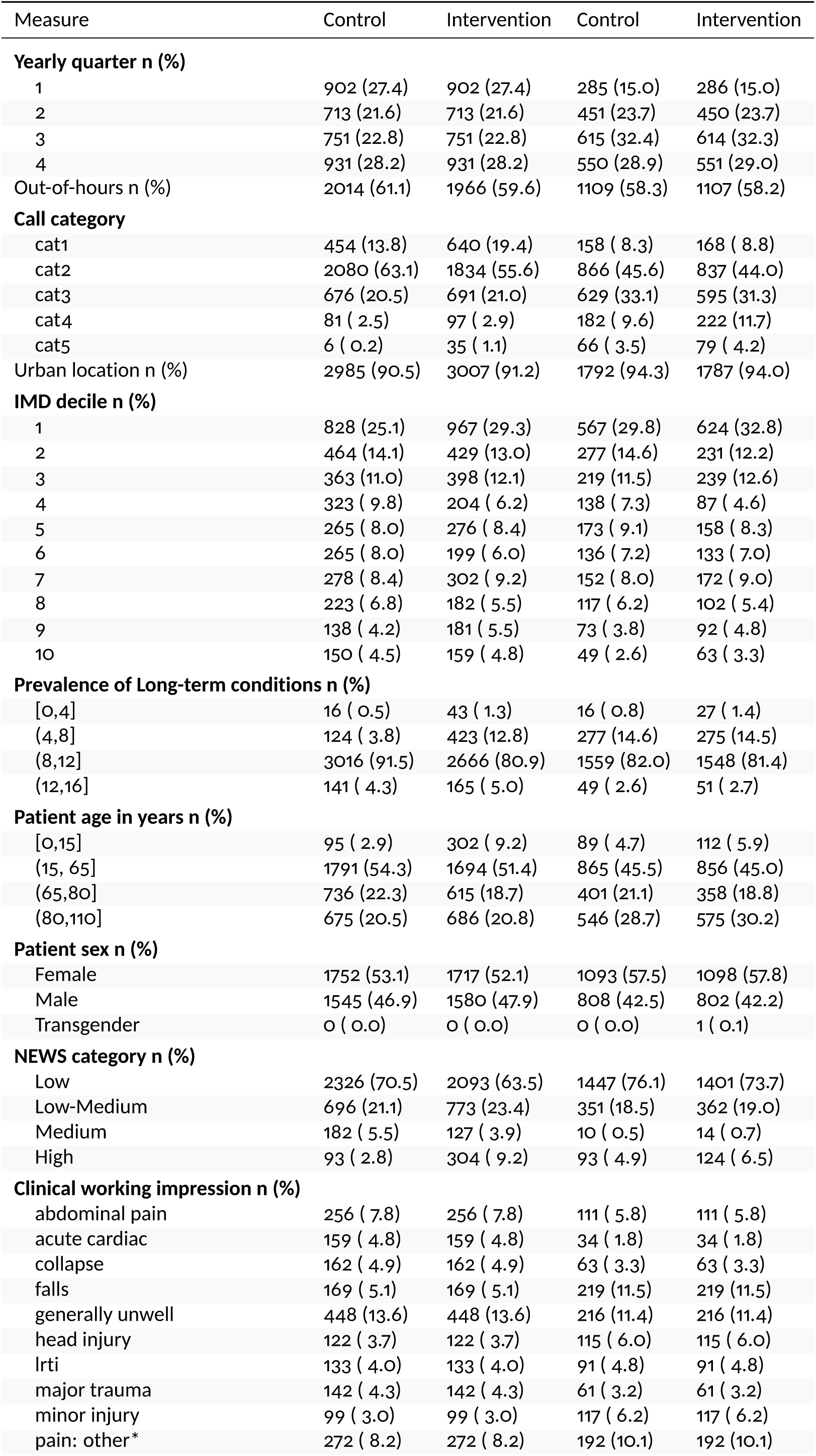
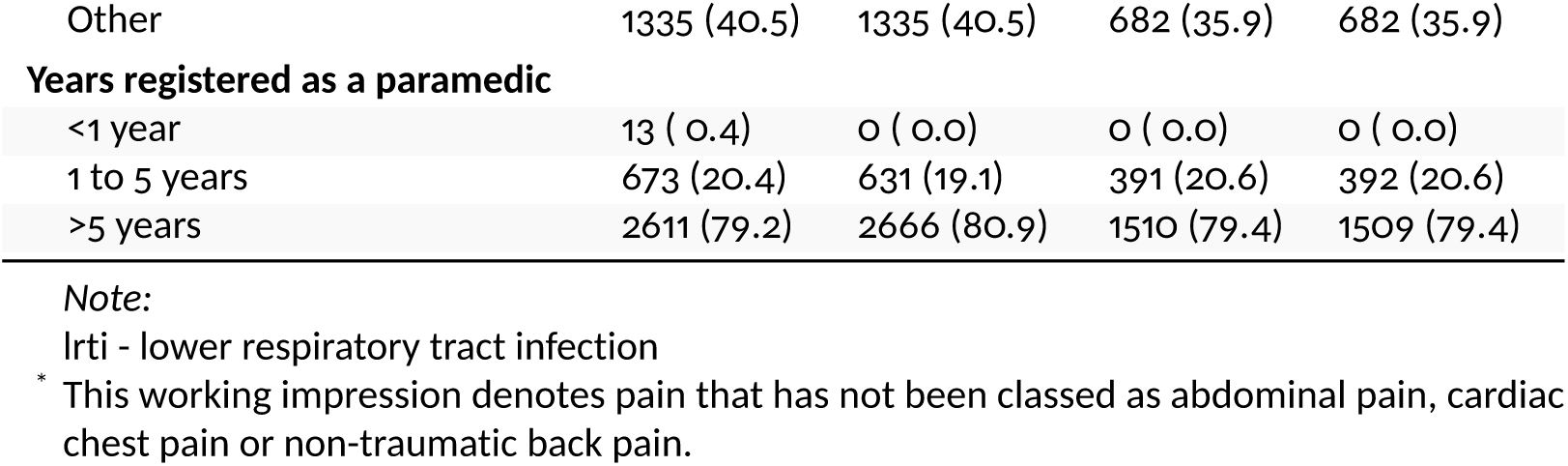
Comparison of matched control and rotational paramedic groups, stratified by pre and post-placement phases

In addition to the substantial reduction in number of cases attended in the post-placement phase, there were also other differences in pre- and post-placement cases, which could have contributed to the change in rate of non-conveyance, validating the decision to include a matched control (Table 1).

### Pre- and Post-rotation exploratory data analysis

Operational activity was lower post-placement since intervention group SPs had to undertake a range of additional activities in the post-placement phase, including staffing a dedicated SP dispatch desk in EOC and working in GP practices as part of the HEE pilot (Table 2). Post-placement, there were also differences in triage call category and physiological acuity based on the NEWS risk category that the SPs were tasked to attend (Supplementary 2).

**Table 2:** Result of segmented regression analysis for appropriate non-conveyance

|  | Coefficient (%) | 95% CI | P value |
| --- | --- | --- | --- |
| Initial control group level | 16.8 | 10.9 to 22.8 | <0.001 |
| Pre-placement control group trend | 0.1 | -0.7 to 0.9 | 0.831 |
| Difference in level between control and intervention groups | 11.8 | 3.4 to 20.2 | 0.007 |
| Intervention group trend relative to control group | -0.3 | -1.4 to 0.9 | 0.622 |
| Post-placement change in control group level | 11.2 | 3.3 to 19.2 | 0.007 |
| Post-placement change in control group trend | 0.1 | -1.0 to 1.2 | 0.862 |
| Post-placement intervention group change in level relative to control group | 35.0 | 23.8 to 46.2 | <0.001 |
| Post-placement intervention group change in trend relative to control group | -1.2 | -2.8 to 0.5 | 0.156 |

Operational activity was higher pre-placement (median 81 (IQR 40–104) hours per month) than post-placement (median 47 (IQR 34–64) hours per month), since intervention group SPs had to undertake a range of additional activities in the post-placement phase, including staffing a dedicated SP dispatch desk in EOC and working in GP practices as part of the HEE pilot. In addition, there were also differences in triage call category (a marker of perceived acuity following telephone triage of the call), and physiological acuity based on the NEWS risk category that the SPs were tasked to attend, (Table 1 and Supplementary 1).

### Time series

Figure 2 illustrates the change in raw and fitted CITS model data between the pre- and post-placement phase. There was no indication of auto-regression, where future values are based on past values (Durbin-Watson statistic 2.37, p=0.79). Post-placement, the intervention group significantly increased their appropriate non-conveyance rate by 35.0% (95%CI 23.8–46.2%, p=<0.001) relative to the control group (Table 2). However, there was a non-statistically significant decrease in the trend of appropriate non-conveyance relative to the control group of −1.2% (95%CI −2.8–0.5%, p=0.156). The sensitivity analysis (excluding working impression as a matching variable) demonstrated a smaller increase appropriate non-conveyance in the intervention group relative to the control group of 27.1% (95%CI 16.4–37.7%, p<0.001), and smaller decrease in the trend of appropriate non-conveyance (−0.9%, 95%CI −2.4–0.6%, p=0.247).

**Figure 2:**
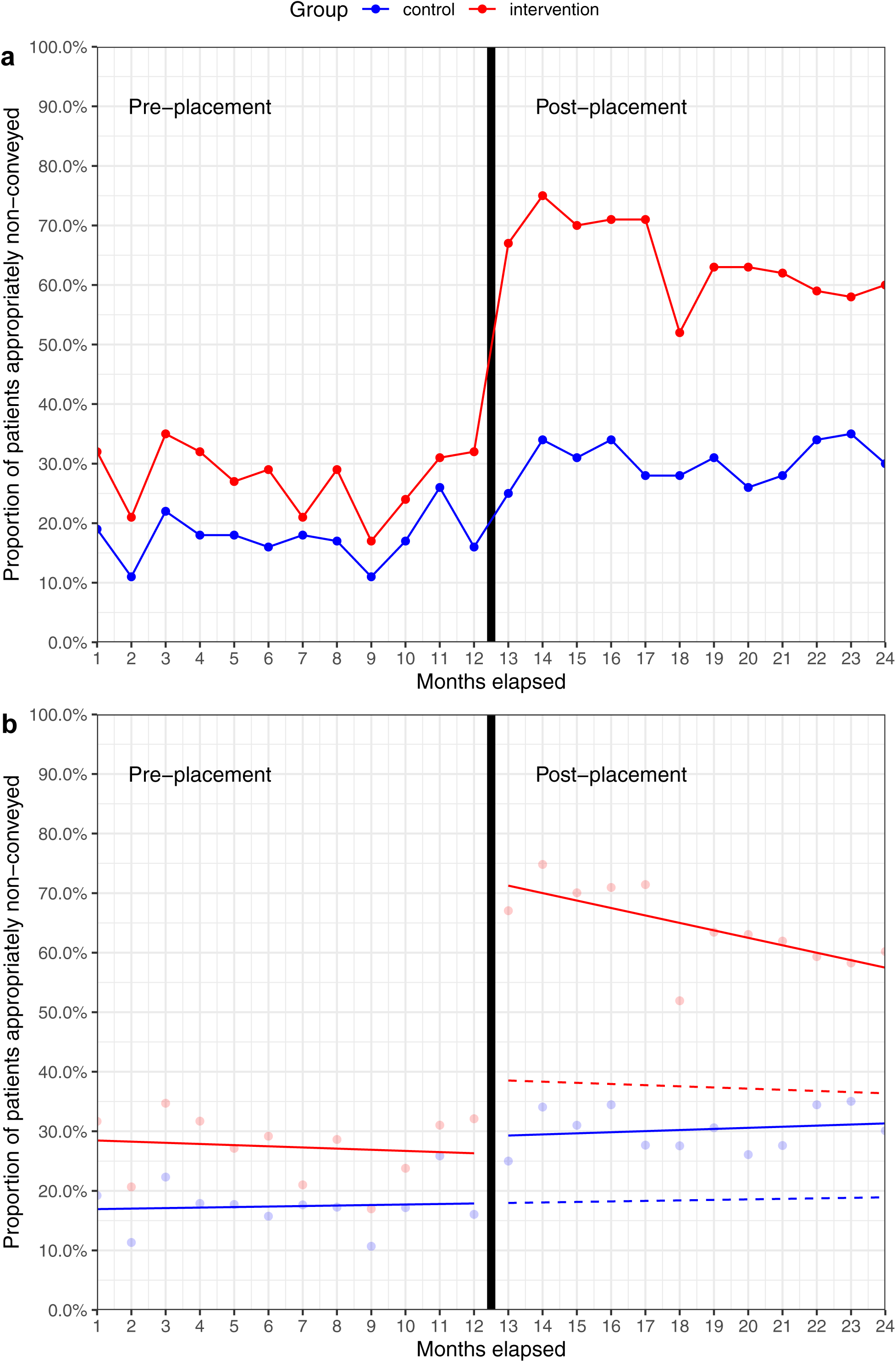
Effect of 10-week primary care placement on appropriate non-conveyance. a) Monthly appropriate non-conveyance rates b) Fitted CITS model. Dashed lines represent the counterfactuals (what would have happened in the absence of the intervention). £735.83 per appropriate non-conveyance (95% bootstrapped CI £709.91–£761.91) and a cost-effectiveness ratio of £2719.1 per percentage increase in appropriate non-conveyance (95% bootstrapped CI £2623.31–£2815.45).

### Economic analysis

The sensitivity analysis (excluding the working impression) calculated the mean post-placement cost per appropriate non-conveyance for intervention group SPs to be £528.24 (95% bootstrapped CI £480.67–£576.22) versus £1264.07 (95% bootstrapped CI £1242.58–£1286.13) for the same group in the pre-placement phase. This represents a mean saving of

## Discussion

In this single NHS ambulance service study, we found a clinically important and statistical significant increase in appropriate non-conveyance of patients following a 10-week GP practice placement. In addition, this intervention proved to be cost saving compared to usual care. These results need to be interpreted with caution, since they only include data from a single ambulance service with less than 10% of all paramedics currently in the role of SP. Training and experiential opportunities do vary between organisations, in part likely due to the piecemeal way in which the advanced practice roles have evolved for paramedics in the UK [9]. YAS has been commissioning the education of SPs with local Higher Education Institutions since 2014, with a focus on minor illness and injury, and long-term conditions, which is likely to resonate with other similar schemes [16]. Furthermore, YAS have continued to work to develop their workforce model aligned to the College of Paramedics post-registration career framework since 2015 [17]. This has placed YAS in a strong position following the introduction of national guidance in 2018 that advocated enhanced training and development for paramedics specifically to reduce the need for hospital admission [18].

There were differences in the acuity and working impressions of patients between pre- and post-GP placement, supporting the choice of methodology for this study. Since cases were matched, it is possible to see how the control group of paramedics also achieved increased rates of appropriate non-conveyance when tasked to cases allocated a lower triage call category and NEWS risk category (Table 1). However, even when accounting for case matching, intervention group SPs had a 35% improvement in appropriate non-conveyance compared to the control group. In addition, there were lower proportions of certain types of working impressions, such as acute cardiac, and higher proportions for others, such as falls and minor injuries, reflecting dedicated tasking by SPs staffing a dispatch desk in EOC. It is possible that the results would have shown a greater difference in proportions of certain presentations pre- and post-placement, had the desk not closed in June 2019.

Overall, SPs attended less cases in the post-placement phase. In addition to attending 999 calls, intervention group SPs also fulfilled other roles, including rotating back into GP surgeries (18% of post-placement hours) and staffing the desk in EOC (29.9%). This resulted in a drop in time responding to 999 calls from 84.4% pre-placement to 60.6% after (Table **??**). While availability for operational shifts reduced, the improved appropriate non-conveyance rates suggest that ambulance services should focus on EOC processes to maximise appropriate dispatching for specialist and advanced practice roles to 999 calls.

The non-conveyance rates seen in this study are difficult to compare with other reported statistics, since the population included in this study is different to all emergency call activity. For example, in the intervention group’s pre-placement phase, there was a higher proportion of category 1 and 2 calls (75–76.9%) compared to YAS figures reported nationally (64%), but a lower proportion post-placement, due to greater case selection with the introduction of the dedicated SP tasking desk [19]. During the study period, YAS ‘see and treat’ rates were between 22.9–25.4% which was lower than the English average of 29.3–30.7% [20].

An evaluation of the first phase of the rotating paramedic pilot reported non-conveyance rates of at least 70%, which mirrors the performance of rotating advanced paramedics in Wales [21]. Two sites in the rotating paramedic pilot had non-conveyance rates in excess of 90%, however these schemes were primary care focused, rather than fully ambulance service based, highlighting the different models commissioned during the pilot. Further evaluation is required to understand the most appropriate model for a paramedic rotation that benefits all parts of the system.

In YAS, the integration into the primary care teams during the Leeds rotation enabled the SPs to develop a greater understanding of the local healthcare system as they navigated pathways across community and acute care. This knowledge could then be utilised when the SPs rotated back into YAS and either responding to 999 calls or working in the EOC to identify appropriate 999 calls for an SP response. However, the impact of improved clinical knowledge and greater understanding of local pathways, and their effect on clinical practice and decision-making is uncertain, and requires further research. Despite this, the value of paramedics being afforded the opportunity to undertake a primary care placement has been demonstrated in this study and supports the qualitative findings from the HEE evaluation. This suggests that support and education from GPs, an appreciation of primary care and other health and social care agencies and the opportunity to develop inter-service, multi-disciplinary relationships across the health and social care system, are beneficial to patient care [10].

### Limitations

We used routine observational data rather than conducting a randomised-controlled trial for example, which was not possible since the rotation had already completed when this study was undertaken. The outcome of this study, while patient focused, could not capture episodes where patients presented to other sectors of the healthcare system. In addition, identifying re-contacts, relied on identification of cases either by NHS number or a combination of patient name, age and incident location which may have been missing on subsequent calls.

We had limited data on the SPs themselves, meaning that it was not possible to determine whether the SPs in the pilot were representative of all YAS SPs, although we match on length of time registered as a paramedic.

The number of missing working impression codes was not anticipated, and so no contingency was made in the methodology to account for this. While the sensitivity analysis showed that this is likely to have had a modest impact on our findings, in retrospect this study would have been more robust with a plan to take account of this.

Finally, it became apparent once the data was provided that determining the grade of paramedic in the control group with certainty was not possible, which may have been a confounding factor as non-pilot SPs may have ended up in the control group. If this is the case, then the results we present here are a conservative estimate of the GP placement and it may in fact, be even more effective at improving safe non-conveyance in a cost-effective manner.

## Conclusion

In this single UK NHS ambulance service study, we found a clinically important and statistically significant increase in appropriate non-conveyance rates by specialist paramedics who had completed a 10-week GP rotation. This improvement persisted for the 12-month period following the rotation and demonstrated cost savings compared to usual care.

## Data Availability

Permission has not been provided to make the original dataset available. However the scripts to undertake the analysis and a synthetic data set are available from the study GitHub repository: https://github.com/RichardPilbery/SPRAINED.

## Acknowledgements

This work uses data provided by patients and collected by the NHS as part of their care and support. The authors would also like to thank the Yorkshire Ambulance service business intelligence team who collated the data used in this study.

## Contributors

RP, TY and AH conceived and designed the study. RP obtained the research approvals and acts as guarantor for the paper. All authors drafted the manuscript and contributed substantially to its revision.

## Funding

This paper presents independent research by the NIHR Applied Research Collaboration Yorkshire and Humber (ARC YH).

## Disclaimer

The views expressed in this publication are those of the author(s) and not necessarily those of the National Institute for Health Research or the Department of Health and Social Care.

## Supplementary 1

Due to the high number of missing working impressions, we conducted a sensitivity analysis including data that was matched without working impression as a variable. Figure 3 shows the change in proportion of appropriate non-conveyance in this group. The results of the segmented regression can be seen in Table 3 and demonstrate a smaller increase appropriate non-conveyance in the intervention group relative to the control group of 27.1% (95%CI 16.4–37.7%, p<0.001), but smaller decrease in the trend of appropriate non-conveyance (−0.9%, 95%CI −2.4–0.6%, p=0.247).

**Table 3:** Result of segmented regression analysis for appropriate non-conveyance (excluding working impression)

|  | Coefficient (%) | 95% CI | P value |
| --- | --- | --- | --- |
| Initial control group level | 18.2 | 12.5 to 23.8 | <0.001 |
| Pre-rotation control group trend | -0.0 | -0.8 to 0.7 | 0.936 |
| Difference in level between control and intervention groups | 9.9 | 1.9 to 17.8 | 0.017 |
| Rotation group trend relative to control group | -0.0 | -1.1 to 1.1 | 0.984 |
| Post-rotation change in control group level | 14.0 | 6.5 to 21.5 | <0.001 |
| Post-rotation change in control group trend | -0.0 | -1.1 to 1.0 | 0.933 |
| Post-rotation intervention group change in level relative to control group | 27.1 | 16.4 to 37.7 | <0.001 |
| Post-rotation intervention group change in trend relative to control group | -0.9 | -2.4 to 0.6 | 0.247 |

**Figure 3:**
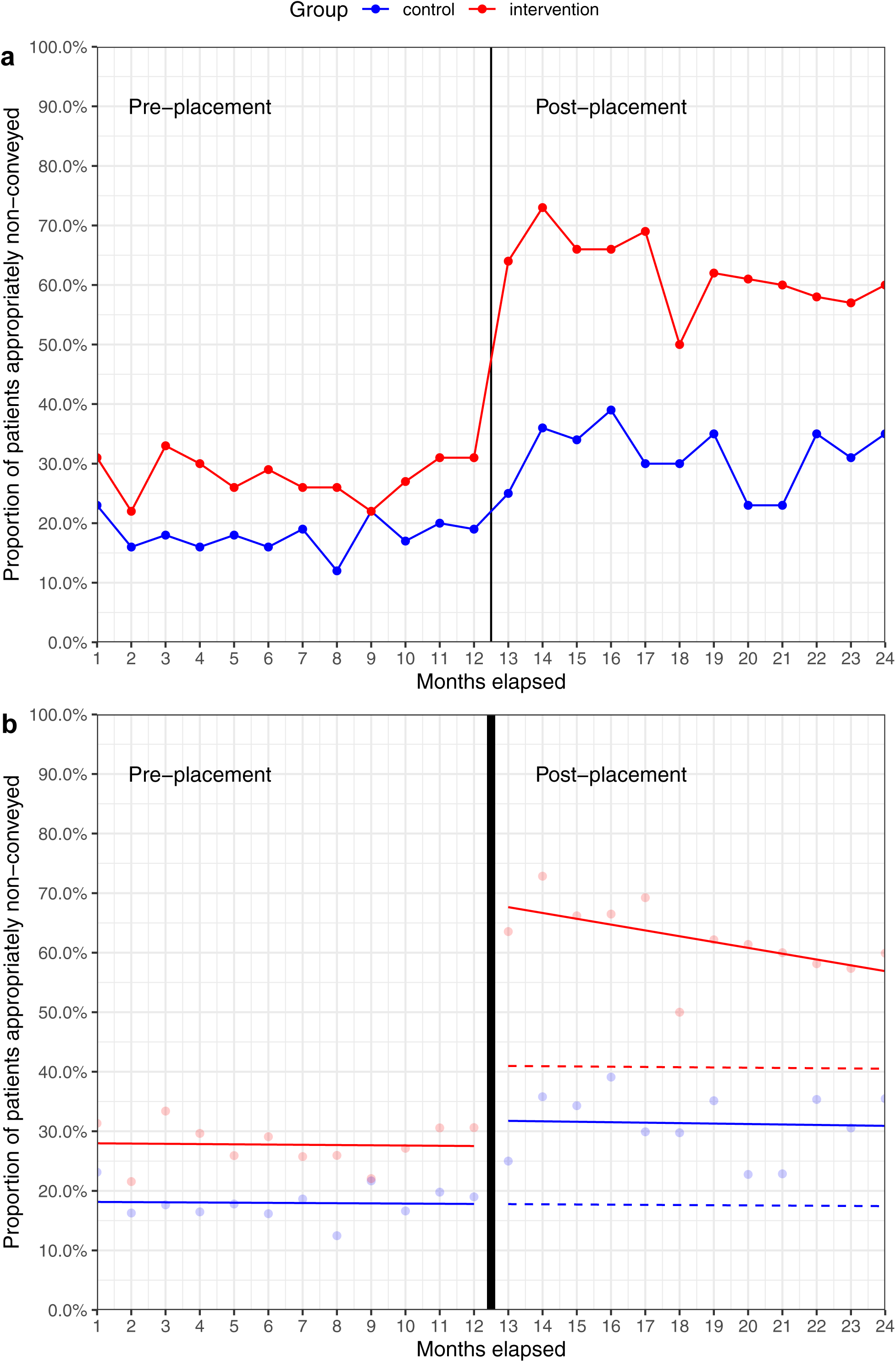
Effect of 10-week primary care rotation on appropriate non-conveyance (excluding working impression). a) Monthly appropriate non-conveyance rates b) Fitted CITS model. Dashed lines represent the counterfactuals.

## Supplementary 2

The following figures highlight the difference in triage call acuity (Figure 4) and NEWS risk category (Figure 5) pre-and post-placement, demonstrating the need for a control in the study design.

**Figure 4:**
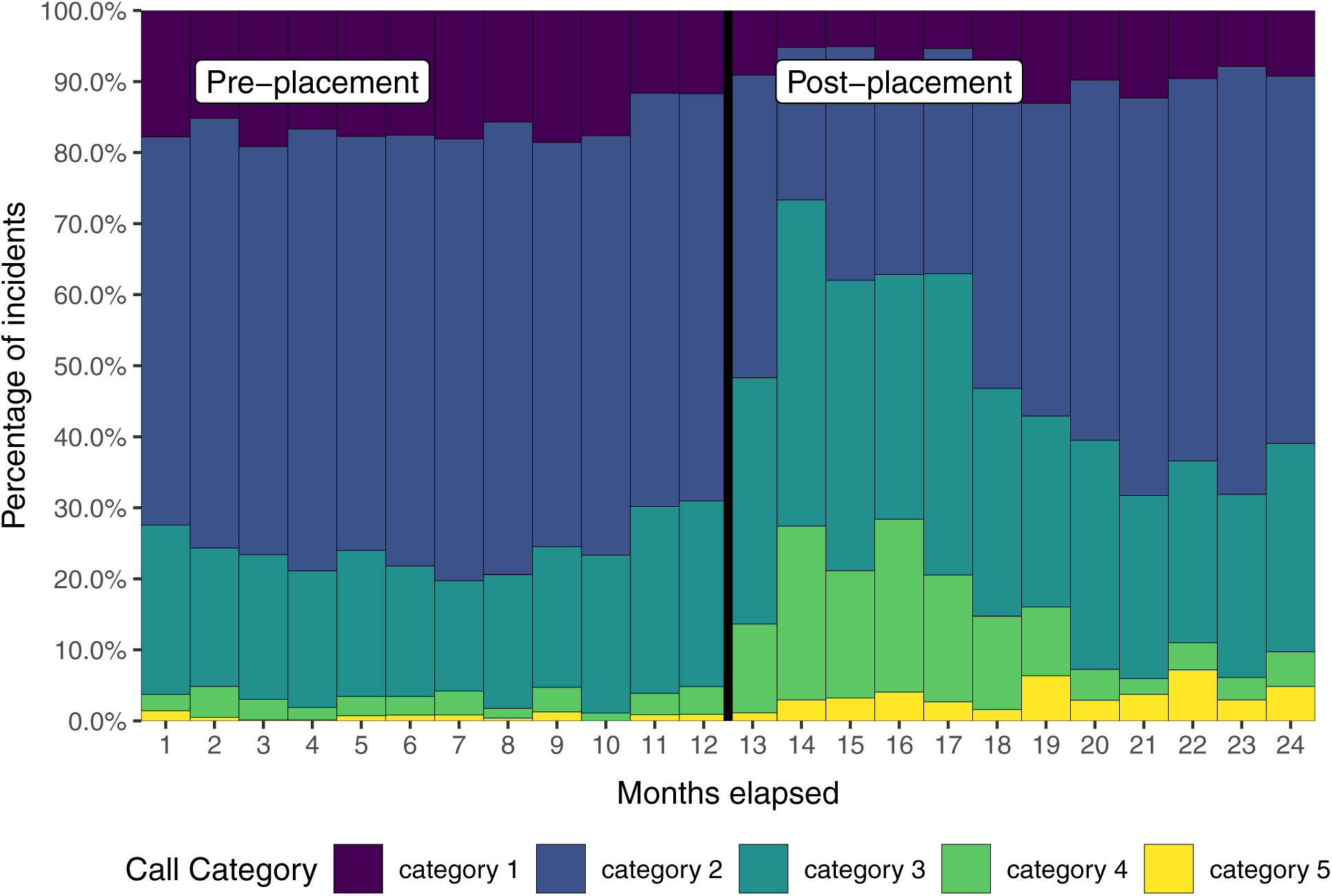
Call category pre- and post-placement

**Figure 5:**
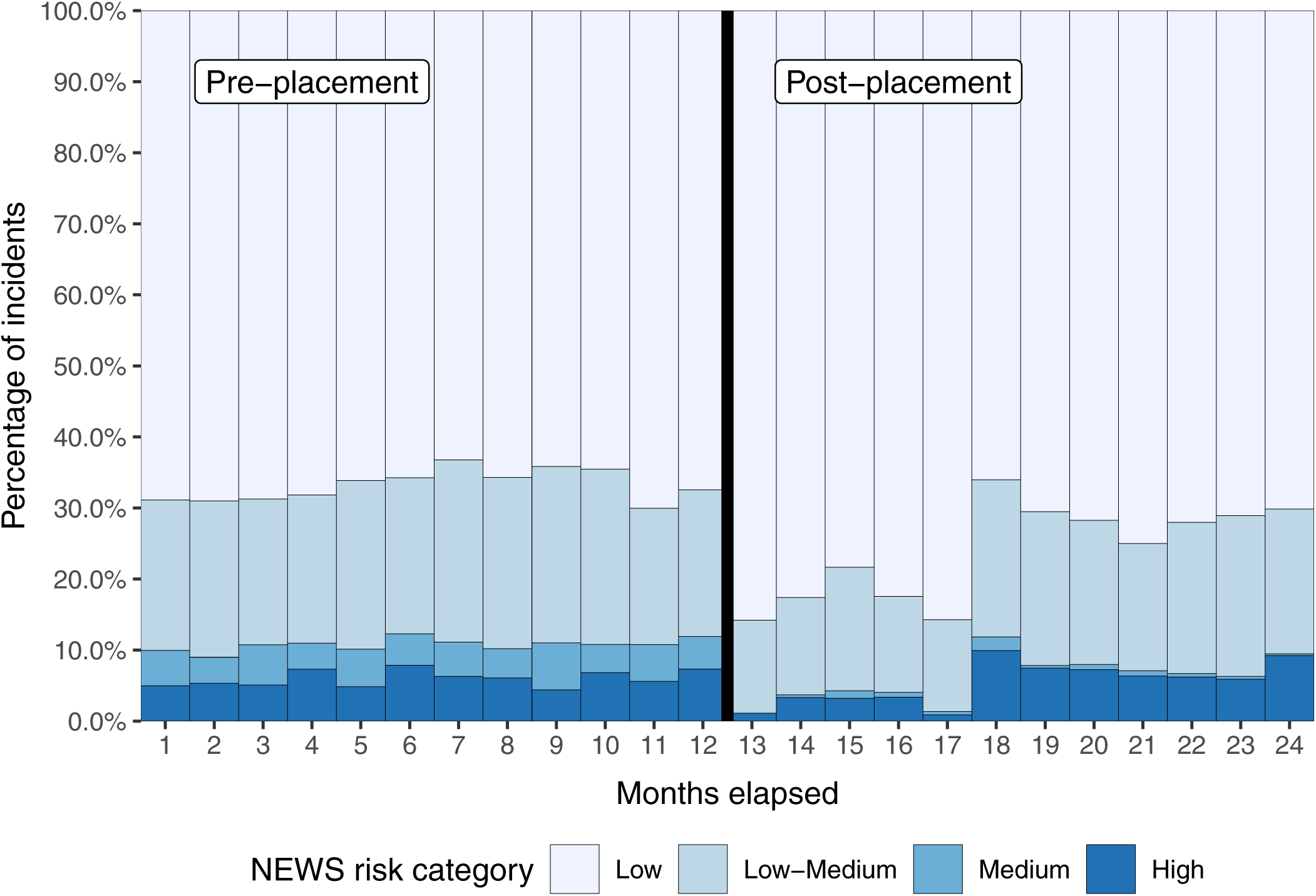
NEWS category pre- and post-placement

## Notes

### Competing Interest Statement

The authors have declared no competing interest.

### Clinical Trial

NCT04193800

### Author Declarations

UK HRA and Health and Care Research Wales (HCRW) approval obtained. REC reference: 20/HRA/0006 IRAS project ID: 265560

### Summary of Updates

Changes in response to reviewer's comments.

